# COVID-19 vaccine acceptance and its socio-demographic and emotional determinants: a multi-country cross-sectional study

**DOI:** 10.1101/2021.05.30.21258074

**Authors:** A. de Figueiredo, C. Simas, H. J. Larson

## Abstract

**Background:** Multiple COVID-19 vaccines have now been licensed for human use, with other candidate vaccines in different stages of development. Effective and safe vaccines against COVID-19 are essential to achieve global control of the pandemic caused by severe acute respiratory coronavirus 2 (SARS-CoV-2), but multiple factors, including vaccine supply and vaccine confidence, will be key for high rates of global uptake. Confidence in COVID-19 vaccines, socio-demographic status, and recent emotional status are likely to be key drivers of COVID-19 vaccine acceptance. In this study, we explore these determinants of COVID-19 vaccination intent across17 countries worldwide.

**Methods:** In this large-scale multi-country study, we explore intent to accept a COVID-19 vaccine and the socio-demographic and emotional determinants of uptake for 17 countries and over 19,000 individuals surveyed in June and July 2020 via nationally representative samples. We used Bayesian ordinal logistic regressions to probe the relationship between intent to accept a COVID-19 vaccine and individuals’ socio-demographic status, their confidence in COVID-19 vaccines, and their recent emotional status. Gibbs sampling was used for Bayesian model inference, with 95% Bayesian highest posterior density intervals used to capture uncertainty.

**Findings:** Intent to accept a COVID-19 vaccine is highest in India, where 77.8% (95% HPD, 75.5 to 80.0%) of respondents strongly agreeing that they would take a new COVID-19 vaccine if it were available. The Democratic Republic of Congo (15.5%, 12.2 to 18.6%) and France (26.4%, 23.7 to 29.2%) have the lowest share of respondents who strongly agree that they would accept a COVID-19. Confidence in the safety, importance, and effectiveness of COVID-19 vaccines are the most widely informative determinants of vaccination intent. Socio-demographic and emotional determinants played a lesser role, with being male and having higher education was associated with increased uptake intent in five countries and being fearful of catching COVID-19 also a strong determinant of uptake intent.

**Interpretation:** Barriers to COVID-19 vaccine acceptance will be highly country and context dependent. These findings highlight the importance of regular monitoring of COVID-19 vaccine confidence to identify groups less likely to vaccinate and to monitor the impact of vaccination policies on uptake behaviour.

## Introduction

Numerous COVID-19 vaccines have now been licensed for human use, with other candidate vaccines in different stages of clinical development^1^. Effective and safe vaccines against COVID-19 are essential to achieve global control of the pandemic caused by severe acute respiratory coronavirus 2 (SARS-CoV-2). Immunisation against COVID-19 can substantially reduce hospitalisations and severe disease^2,3^, and – if administered to and accepted by sufficient fractions of the population – achieve herd immunity. Vaccination will be critical in reducing excess mortality and morbidity, as well as relieving economic and societal burdens associated with COVID-19.

Successful roll out of COVID-19 vaccines will depend on logistic aspects (e.g., at-scale manufacture, fast and equitable distribution) and also on global uptake of COVID-19 vaccines. To bring the pandemic under control, local communities will need to accept vaccination. Public acceptance of vaccines is dependent on public confidence in the safety, importance, and effectiveness of the vaccine. Past experiences with specific vaccines or the health systems more generally, previous vaccine controversies, and exposure to mis- or disinformation can undermine confidence in vaccination. Vaccine confidence is highly context dependent and can vary markedly between and within countries^4–6^. Recent surveys quantifying COVID-19 vaccine acceptance have indicated wavering willingness to vaccinate globally^6,7^. Studies have identified barriers to uptake, such as anxieties around the speed of vaccine development and fears over safety and relaxation of regulatory rules^8^, fears over the use of new vaccine technologies such as mRNA, and misinformation circulating on social media^9,10^. Emotions also influence COVID-19 vaccine acceptance(hope) or hesitancy(fear). Healthcare systems in some settings also face public, and even health care professional, reluctance to vaccinate. Fears around vaccine side-effects or over contracting COVID-19 while going to a health facility to receive a vaccine, can drive hesitancy, while misinformation or unregulated social media platforms may seek to exploit these fears.

In this large-scale multi-country study, we explore intent to accept a COVID-19 vaccine and the socio-econo-demographic and attitudinal barriers to acceptance across 17 countries and over 19,000 individuals. The countries surveyed were selected to represent a range of countries in different regions, with varying economic and political contexts. to evaluate COVID-19 vaccine acceptance. A range of putative drivers of COVID-19 vaccine acceptance are considered and include socio-econo-demographic characteristics (sex, age, highest educational attainment, work status, and religious affiliation); confidence in the safety, importance, and effectiveness of a COVID-19 vaccine; and emotional drivers, such as fears and anxieties about COVID-19. In addition, to understand population-specific concerns about accepting a COVID-19 vaccine, specific reasons for non-acceptance are explored. Our findings are discussed in light of vaccination policy and historic challenges surrounding vaccine confidence.

## Methods

### Data

A total of 19,243 individuals (aged 18 and over) are surveyed across 17 countries: Argentina, Brazil, DRC, Ecuador, Ethiopia, France, Germany, India, Italy, South Korea, Lebanon, Nigeria, Pakistan, Peru, Saudi Arabia, the United Kingdom (UK), and the United States of America (USA) (figure 1). The number of respondents ranges from 500 (Democratic Republic of Congo) to 2,500 (USA), with a median of 1,000 and mean of 1,132 respondents. The survey was conducted online in six countries (France, Germany, Italy, South Korea, UK, and USA), face-to-face in Nigeria, and using a computer-assisted telephone interview (CATI) methodology in 10 (Argentina, Brazil, Democratic Republic of Congo, Ecuador, Ethiopia, India, Lebanon, Pakistan, Peru, and Saudi Arabia). Fieldwork was conducted in June and July 2020 (see appendix table A2). Respondents were sampled to match proportions of national demographic breakdowns for sex, age, and sub-national region. Survey weights account for mismatches between these expected distributions and those obtained via the sampling methodologies.

**Figure 1.**
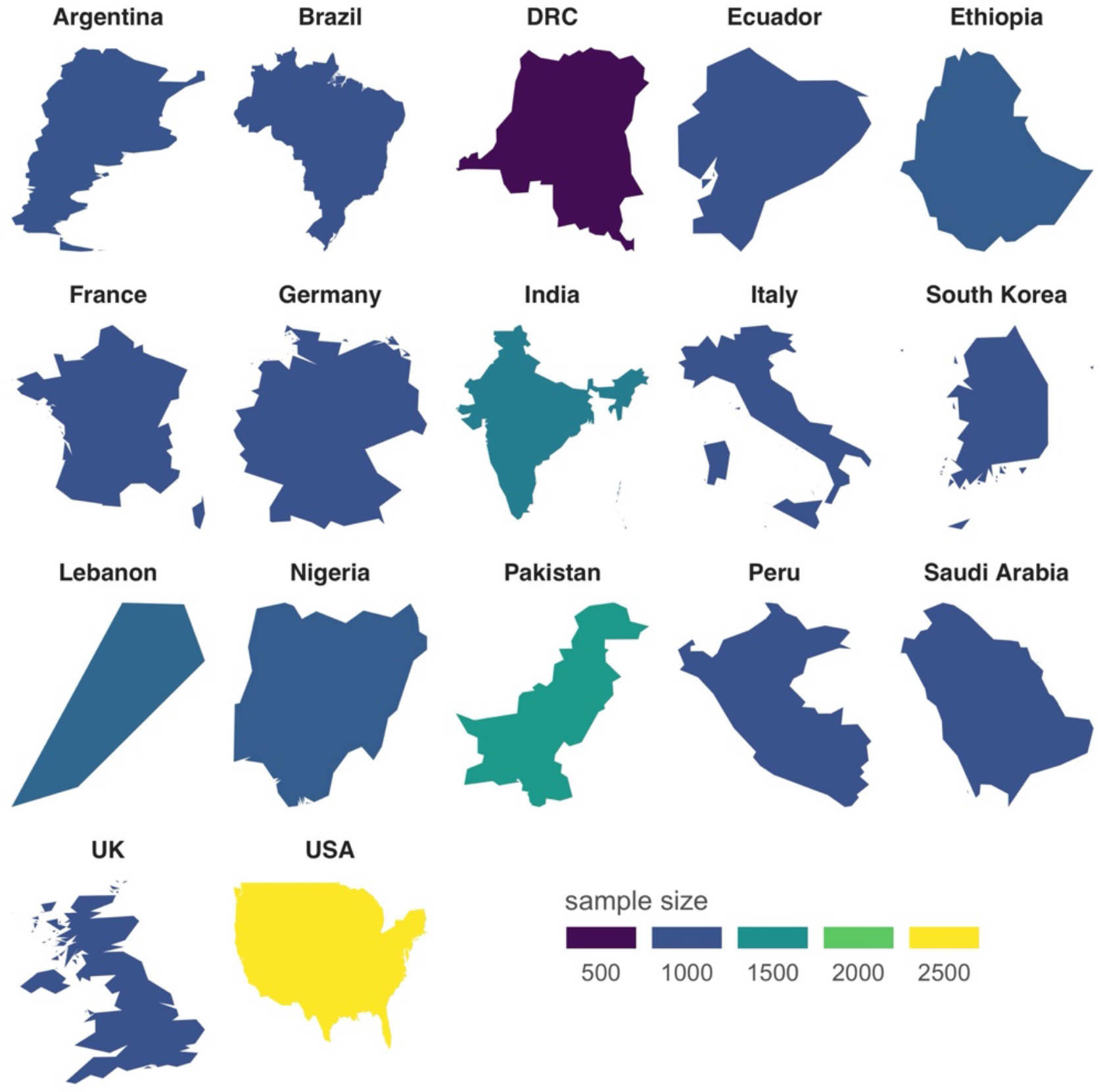
Study settings. Nationally representative surveys are conducted in 17 countries worldwide

#### Response variable

Respondents are asked to rate the extent to which they agreed that they would accept a COVID-19 vaccine if it became publicly available (“*If a new coronavirus (COVID-19) vaccine became publicly available, I would take it*”). Responses were collected on a five-point scale: “*strongly agree*”, “*agree*”, “*do not know*”, “*disagree*”, and “*strongly disagree*”.

#### Covariates

A number of additional variables are collected for each respondent and are used to assess the relationship between the response variable and a) socio-demographic status, b) confidence in a COVID-19 vaccine, c) factors relating to COVID-19, such as whether a respondent is in an at-risk group or if they know anybody who has contracted the disease, and d) emotional determinants. Descriptions for all variables used in the study are provided in table 1. Cross-tabulations of socio-demographic breakdowns by response variable are provided for each country in Table A1 in the appendix. These covariate data were selected from a larger set of possible determinants of uptake from a larger questionnaire that included items on, for example, sources of trust for information about COVID-19, a broader suite of recent emotions including boredom, fear, positivity, etc, and COVID-19 hygiene behaviours such as mask use and handwashing. The covariates selected for this study were those anticipated to have the strongest association with intent to vaccinate, such as confidence about a COVID-19 vaccine, socio-demographics (which may be direct targets for intervention), and aversive emotions. All other questionnaire items were not used during this study once initially discarded. The full questionnaire is provided in the appendix.

**Table 1.** Study data. Outline of all data used throughout this study. The survey items are shown with the possible responses (including recodes, if any), and baselines used in the multivariate ordinal logistic regressions (provided for explanatory variables). COVID-19 vaccination intent is the study response variable. The explanatory factors include socio-econo-demographics, COVID-19 vaccine confidence, COVID-19 questions, and emotional determinants.

| Survey question | Values (recode in parenthesis) | Baseline for regressions |
| --- | --- | --- |
| **COVID-19 vaccination intent (response)** |  |  |
| If a new coronavirus (COVID-19) vaccine became publicly available, I would take it | strongly agree (5), agree (4), do not know (3), disagree (2), strongly disagree (1) | n/a |
| **socio-econo-demographic (SED)** |  |  |
| sex | male and female | male |
| age | 18-24, 25-34, 35-44, 45-54, 55-64, 65+ | 18-24 |
| highest educational attainment | none or no formal education (none/other), primary, secondary, university (undergraduate), master/PhD (postgraduate), other educational level (none/other) | secondary |
| work status | full-time, part-time, unemployed, student, housewife, retired/disabled, refused or do not know or did not answer (prefer not to say) | full-time |
| religious affiliation | atheist/agnostic, Buddhist, Muslim, other Christian, other religion, Protestant, Roman Catholic, Russian or Eastern Orthodox, refused or do not know or did not answer (prefer not to say) | Roman Catholic except for Pakistan and Saudi Arabia (Muslim).<sup>1</sup> |
| Children under 18 living in the household | none (no), children of 0-2 (yes), 3-6 years (yes), 7-12 (yes), or 13-17 years (yes) | no |
| **confidence in a COVID-19 vaccine** |  |  |
| I think a new coronavirus (COVID-19) vaccine would be important | strongly agree (agree), agree, do not know (disagree), disagree, strongly disagree (disagree) | disagree |
| I think a new coronavirus (COVID-19) vaccine would be safe |  |  |
| *I think a new coronavirus (COVID-19) vaccine would be effective* |  |  |
| **COVID-19 battery** |  |  |
| how well informed do you feel you are about coronavirus (COVID-19)? | very well informed (well), fairly well informed (well), not very well informed (not well), not informed at all (not well), I have never heard of [COVID-19] | well |
| do you personally know anyone who has tested positive for COVID-19? If yes, was that a family member, work colleague, friend, or someone else? | no, yes – myself, yes – family member in my household, yes – family member outside my household (yes), yes – a work colleague (yes), yes – a friend (yes), yes – someone else (yes) | no |
| are you taking any non-prescribed medicines or treatments that you have read/heard about that are said to help protect yourself specifically against coronavirus (COVID-19)? | yes, no | no |
| **emotional determinants** |  |  |
| I am afraid that either myself or someone in my household may catch coronavirus (COVID-19) | strongly agree (agree), agree, do not know (disagree), disagree, strongly disagree (disagree) | disagree |
| have you experienced anxiety in the last few days? | yes, no | no |
| have you experienced stress in the last few days? | yes, no | no |
| have you experienced anger in the last few days? | yes, no | no |
| have you experienced fear in the last few days? | yes, no<sup>2</sup> | no |

<sup>1</sup> No religious affiliation data were collected for respondents in India.
<sup>2</sup> These questions were taken from a larger battery of emotional determinants. The factors most likely

### Statistical methods

National-level estimates of intent to accept a COVID-19 vaccine are obtained via posterior samples from a multinomial distribution *y∼Multi*(***p***, *n*) with an uninformative Dirichlet prior over model probabilities, *p∼Dir*(1,1,1,1,1). *y=(y*_*sa*_, *y*_*a*_, *y*_*dk*_, *y*_*d*_, *y*_*sd*_) is the (weighted) count of responses falling into each of the five possible responses and *n* = ∑_k_ y_k_ where *k* ∈ {*sa,a, dk, d, sd*} (*sa* = strongly agree, *a* = agree, *dk* = do not know, *d* = disagree, *sd* = strongly disagree).

Univariate Bayesian linear regressions are used to quantify the association between national-level intent to accept a COVID-19 vaccine and national-level vaccine confidence and the Bayesian R-squared^11^ is used to calculate the strength of association.

Bayesian ordinal logistic regressions are used to explore the link between intent to accept a COVID-19 vaccine and the set of explanatory variables via a multiple regression for each country (see table 1 for model covariate definitions). The outcome variable – intent to accept a COVID-19 vaccine – is given an ordinal scale so that “strongly agree” = 5 and “strongly disagree” = 1. Gibbs sampling is used to estimate the posterior distribution of model parameters using 50,000 samples following model burn-in. The Bayes factor (BF) is used to assess the fit of each of the 17 regressions by comparing each model’s marginal likelihood with that model’s respective null model (an intercept-only model). Bayes factors are computed via Monte Carlo simulation. In each case, it is found that the log Bayes factor greatly exceeds two for each model, providing “decisive” support for each full model over its respective null^12^.

Relevant statistics for parameters of interest (percentages, odds-ratios and log odds-ratios) are reported as a mean estimate (the effect size) with a corresponding 95% highest posterior density (HPD) credible interval. Throughout the study, we remark on log odds ratios if the 95% HPD interval excludes zero (or one, in the case of odds ratios).

`R` version 4.0.3 is used for all statistical analyses. JAGS v 4.3.0 is used (via `rjags`) to implement Gibbs sampling

## Results

### COVID-19 vaccine acceptance

Model-based estimates of COVID-19 vaccine acceptance intent are shown in figure 1A. India (77.8%, 95% highest posterior density, HPD, 75.5 to 80.0%) has the highest proportion of respondents strongly agreeing that they would take a new COVID-19 vaccine if it were publicly available (figure 1A). India is followed distantly by Ethiopia (54.3%, 51.5 to 57.4%), and then Nigeria (44.5%, 41.7 to 47.4%), Argentina (44.4%, 41.4 to 47.5%), and Saudi Arabia (44.2%, 41.7 to 46.6%). The Democratic Republic of Congo (DRC, 15.5%, 12.2 to 18.6%) and France (26.4%, 23.7 to 29.2%) have the lowest share of respondents who strongly agreed that they would accept a COVID-19 vaccine if it were publicly available, followed by the USA (29.7%, 27.9 to 31.4%).

Nigeria (13.5%, 11.6 to 15.5%), Pakistan (14.0%, 11.9 to 16.0%), and the DRC (26.9%, 23.2 to 30.9%) have the highest share of respondents who “strongly disagree” that they would take a COVID-19 vaccine if publicly available. South Korea has the lowest share of respondents who “strongly disagree” that they would take a new COVID-19 vaccine (1.1%, 0.5 to 1.7%).

The values from figure 1A are repeated in figure 1B, but with countries ranked by the percentage of respondents who agree (“agree” or “strongly agree”) that they would take a COVID-19 vaccine. India, Ethiopia, Saudi Arabia, South Korea, and Nigeria rank in the top five under this overall agree metric, while DRC, France, USA, Germany, and Italy the bottom five.

#### COVID-19 vaccine intent and vaccine confidence

There is a strong association between national level vaccine confidence and intent to accept a COVID-19 vaccine (figure 2C). Countries with higher proportions of respondents strongly agreeing that a COVID-19 vaccine would be important (Bayesian *R*^2^ = 0.86, 0.69 to 0.97), safe (0.90, 0.78 to 0.97), and effective (0.94, 0.88 to 0.98) have higher proportions strongly agreeing that they would accept a COVID-19 vaccine.

**Figure 2.**
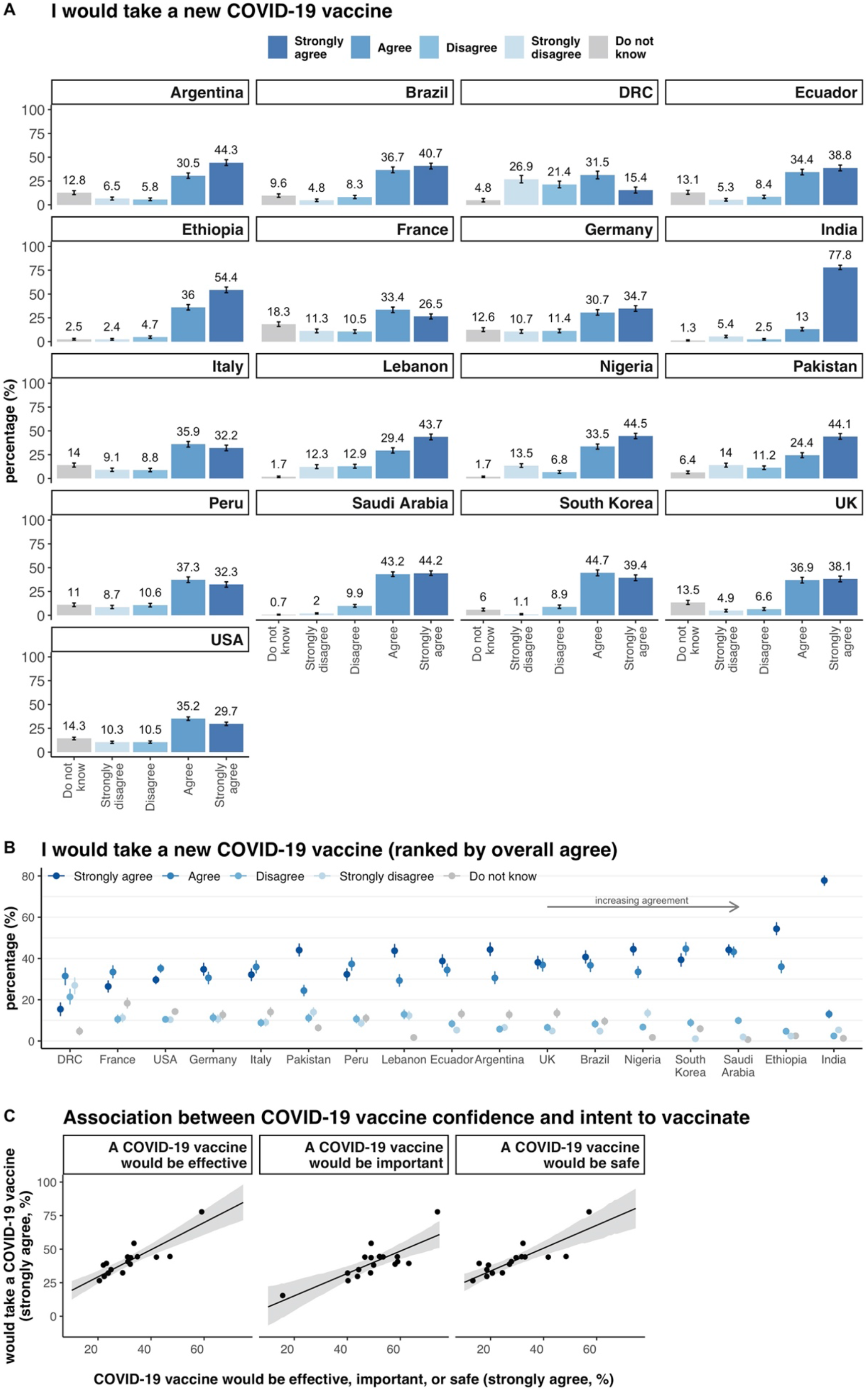
National level trends in intent to accept a COVID-19 vaccine and their links to vaccine confidence. Estimated intent to accept a COVID-19 vaccine for each survey response (A) and ranked by the overall percentage of respondents who agree that they would accept a COVID-19 vaccine (B). The link between national-level confidence in COVID-19 vaccines and intent to accept a COVID-19 vaccine (C).

#### Summary of COVID-19 uptake intent determinants

Figure 3A shows the regression parameters for all 17 multiple regressions, with a summary count of the number of times (across all 17 regressions/countries) that a variable has an odds ratio of association with vaccine uptake intent whose 95% HPD excludes zero in figure 3B. We find confidence in the importance (16 out of 17 countries), safety (16), and effectiveness (all 17) of a novel COVID-19 vaccine are most consistently associated with uptake intent of a COVID-19 vaccine (figure 3B). Sex and emotional characteristics also appear to be strongly connected to uptake intent, with evidence to suggest that five countries have a strong association between individuals’ sex and uptake intent, and a further five that have strong associations between being afraid of catching COVID-19 and uptake intent. In figure 4, the full results of the multiple regressions are shown for each country, including the effect sizes (odds/log odds ratios) and 95% HPD credible intervals (figure 4A-Q). In the following sub-sections, we comment on the effect of each type of explanatory variable (COVID-19 vaccine confidence, socio-econo-demographic, COVID-19-related, and emotional determinants) on vaccine intent.

**Figure 3.**
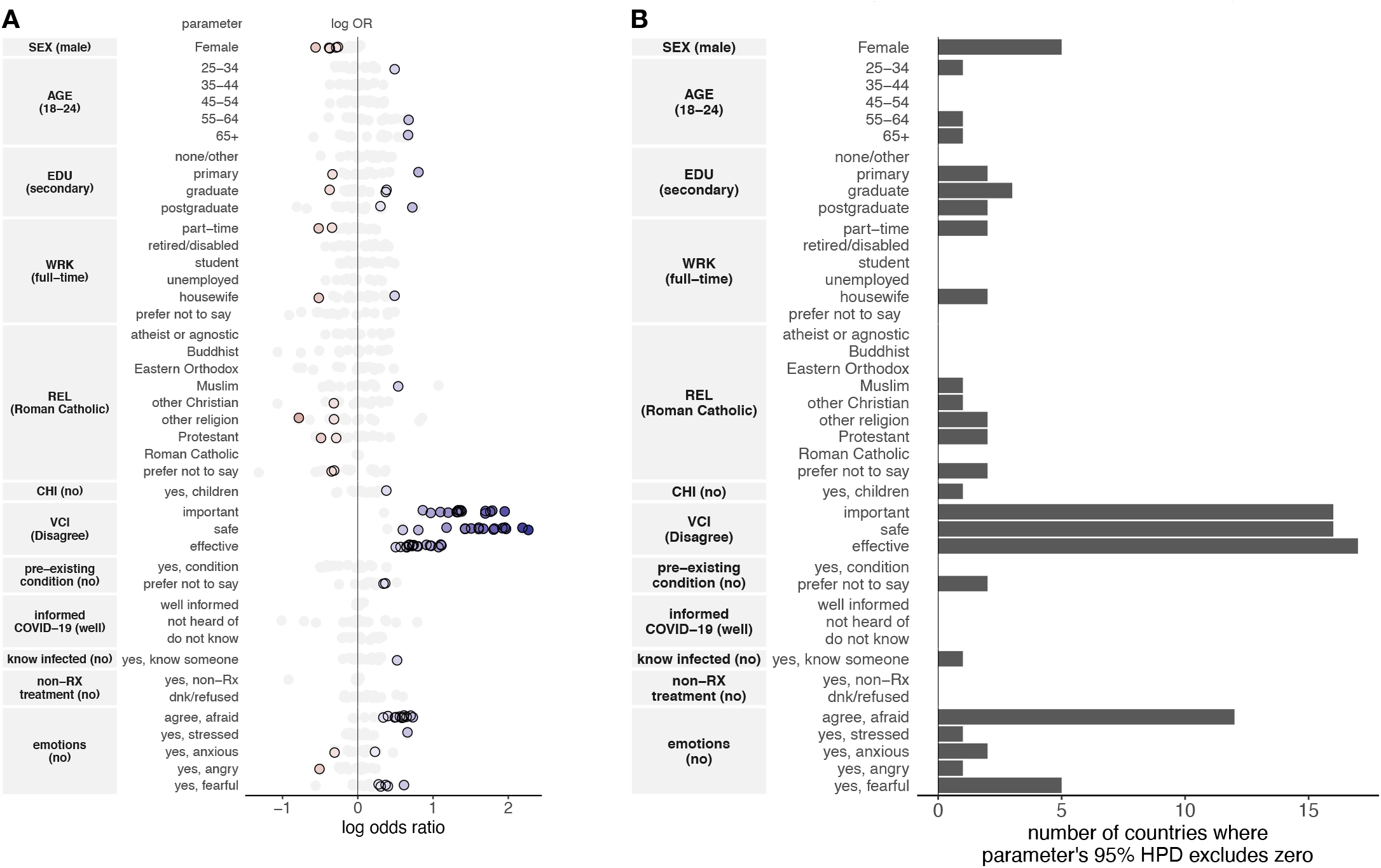
Summary of covariates associated with intent to accept a COVID-19 vaccine. Model parameters across each of the 17 models showing all parameters whose 95% HPD interval excludes zero (A) and a summary count of the number of parameters whose 95% HPD interval excludes zero across all covariates (B).

**Figure 4.**
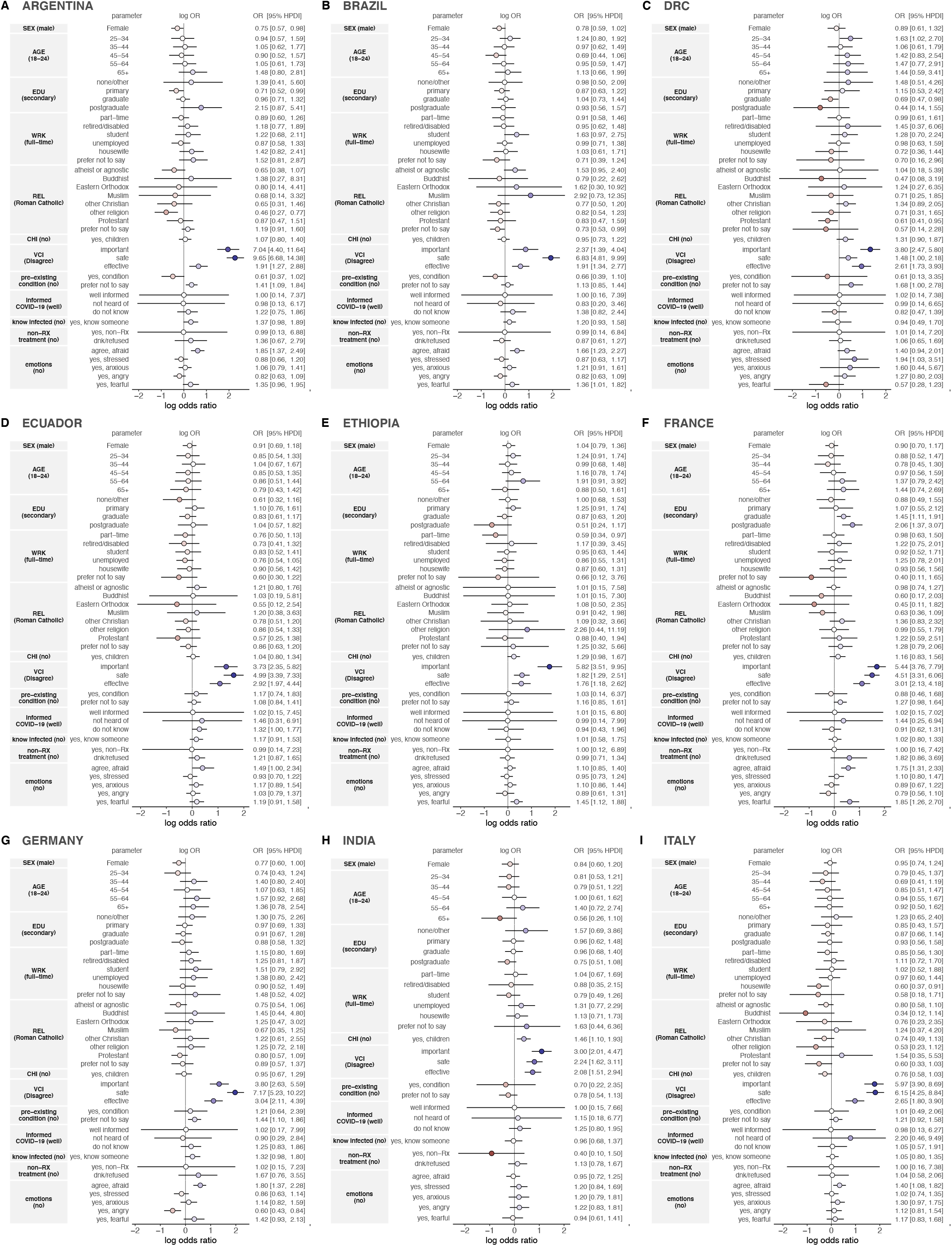
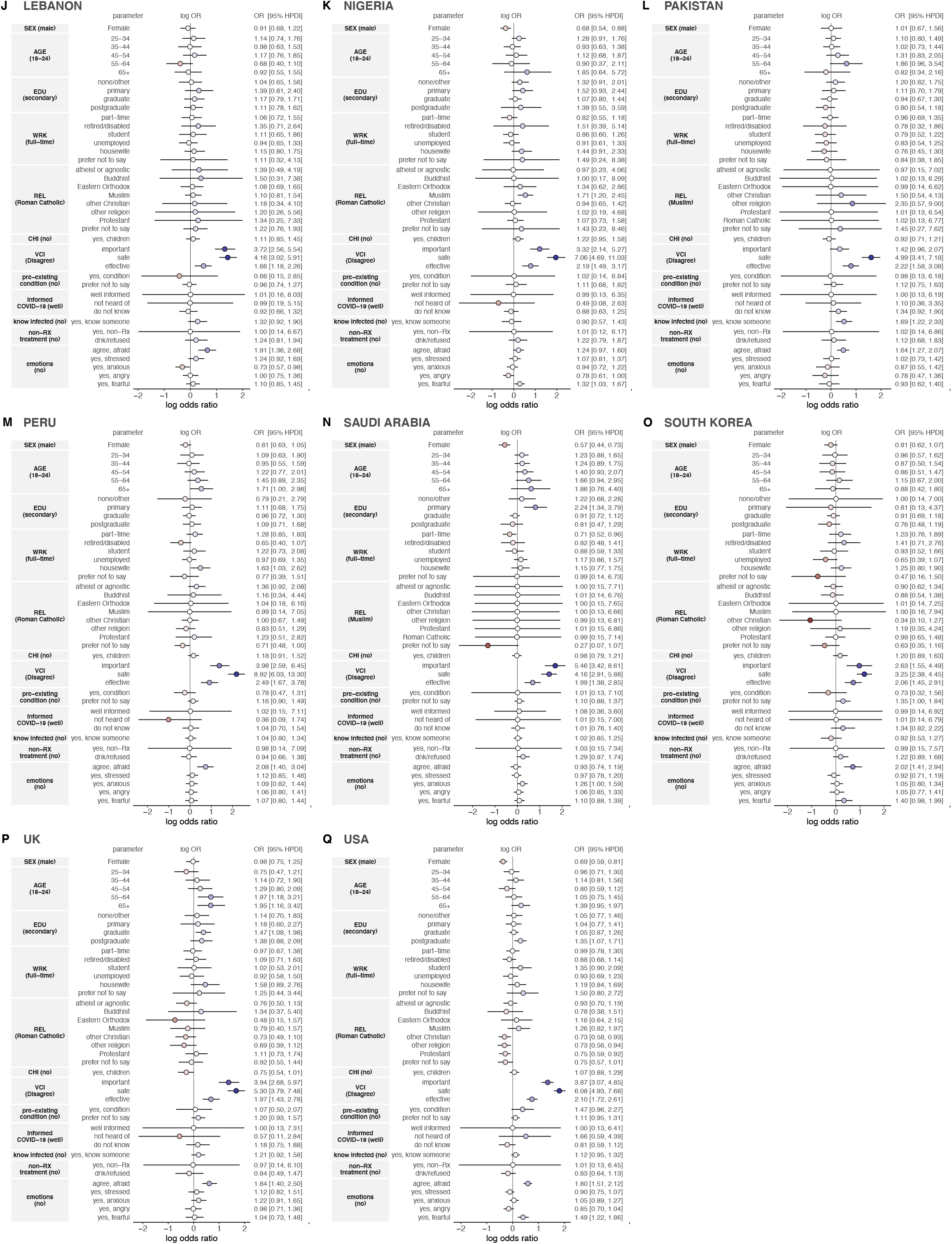
Determinants of intent to accept a COVID-19 vaccine for 17 countries worldwide.

#### Determinants: COVID-19 vaccine confidence

In every country except Pakistan and the DRC (figure 4C and L, respectively), the 95% HPD intervals around inferred odds ratios for all confidence parameters exclude one, revealing that perceptions towards COVID-19 vaccine importance, safety, and effectiveness are all somewhat independently informative of uptake intent. In the DRC, perceptions towards the importance and effectiveness of a novel COVID-19 vaccine appear to be more influential in driving uptake intent than safety perceptions (figure 4C); while in Pakistan, perceptions towards the safety and effectiveness of a vaccine are the most important drivers.

#### Determinants: socio-econo-demographics

Individuals’ sex was informative of uptake intent in five countries (figure 3) and in all these settings females were less likely than males to signify intent to accept a COVID-19 vaccine: Argentina (odds ratio 0.75, 95% highest posterior density interval 0.57 to 0.98), Germany (0.77, 0.60 to 1.00), Nigeria (0.68, 0.54 to 0.88), Saudi Arabia (0.57, 0.44 to 0.73) (where the strongest effect was observed), and USA (0.69, 0.59 to 0.81). (See figures 4A, G, K, N, Q, respectively).

Education is associated with uptake intent in five countries. In Argentina and Saudi Arabia (figures 4A and N, respectively), individuals reporting primary education as their highest educational level are less likely than those with secondary education to agree that they would accept a COVID-19 vaccine (0.71, 0.52 to 0.99 and 2.24, 1.34 to 3.79, respectively). Higher education levels are also found to be associated with increased agreement of vaccine intent in France (where graduates and postgraduates are more likely than those with secondary education to signal intent to accept a COVID-19 vaccine, 1.45, 1.11 to 1.91 and 2.06, 1.37 to 3.07, respectively) and the USA (where postgraduates are more likely, 1.35, 1.07 to 1.71) (figure 4 F and Q, respectively). The DRC (figure 4C) is the only country where we find that those with a higher education level (graduates) are less likely than those with secondary education to agree that they would accept a COVID-19 vaccine (0.69, 0.47 to 0.98).

Other socio-demographic factors were found to play a role in modulating uptake intent, but these factors played less of a consistent role across countries. For example, over 65s in Peru (1.71, 1.00 to 2.98) and the UK (1.95, 1.16 to 3.42) were more likely than 18-24-year-olds to agree they would accept a COVID-19 vaccine (see figure 4 M and P, respectively). (50-64-year-olds were also more likely than 18-24-year-olds in the UK.) Religion was found to be informative of vaccine acceptance intent in Argentina (figure 4A), where other religions were less likely than Roman Catholics to signify intent to accept a COVID-19 vaccine (0.46, 0.27 to 0.77); Brazil (figure 4B), where individuals refusing to provide their religious affiliation were associated with lower uptake intent than Roman Catholics (0.73, 0.53 to 0.99); and Nigeria (figure 4K), where Muslims were more likely than Roman Catholics to intend to take a vaccine (1.71, 1.20 to 2.45). Part-time employment in Ethiopia (0.59, 0.34 to 0.97) and housewives in Italy (0.60, 0.37 to 0.91), were both less likely than those in full-time employment to report intent to accept a vaccine (figure 4E and I respectively). In India (figure 4H), individuals who report not having a child under 18 in the house were more likely to report intending to vaccinate than those who did (1.46, 1.10 to 1.93). There was not enough evidence to suggest that the socio-demographic variables in the study played a role in impacting uptake intent in Ecuador, Lebanon, Pakistan, and South Korea (figure 4 D, J, L, and O, respectively).

#### Determinants: COVID-19

We find evidence to suggest that individuals who “prefer not to say” whether they or somebody in their household has underlying conditions which may increase their risk from COVID-19 are more likely to report (than those who report that there is nobody in their household at risk) that they would take a COVID-19 vaccine in Argentina (1.41, 1.09 to 1.80), DRC (1.68, 1.00 to 2.78), and Germany (1.44, 1.10 to 1.86), see figures 4A, C and G, respectively). If respondents are unwilling to disclose potentially sensitive information about medical conditions of either themselves or their household, then these results could suggest that individuals are more likely to vaccinate themselves to protect other members of their household.

In only one country (Pakistan, figure 4L) is there evidence to suggest that knowing somebody who has been infected by SARS-CoV-2 increases your intent to vaccinate, though this effect is notably strong (1.69, 1.22 to 2.33). We find no evidence to suggest that self-reported awareness about COVID-19 or whether individuals are taking non-prescribed medication to treat or prevent COVID-19 plays a role in uptake intent. Although, we note that 1,941 (10.1%) of respondents surveyed across all countries report taking non-prescribed medicines or treatments to protect themselves against coronavirus (“*are you taking any non-prescribed medicines or treatments that you have read/heard about that are said to help protect yourself specifically against Coronavirus (COVID-19)? By non-prescribed, I mean over the counter medicine, herbal medicine, alternative treatments or supplements”*), with over one in five respondents in South Korea reporting that they are taking non-prescribed treatments (figure 5).

**Figure 5.**
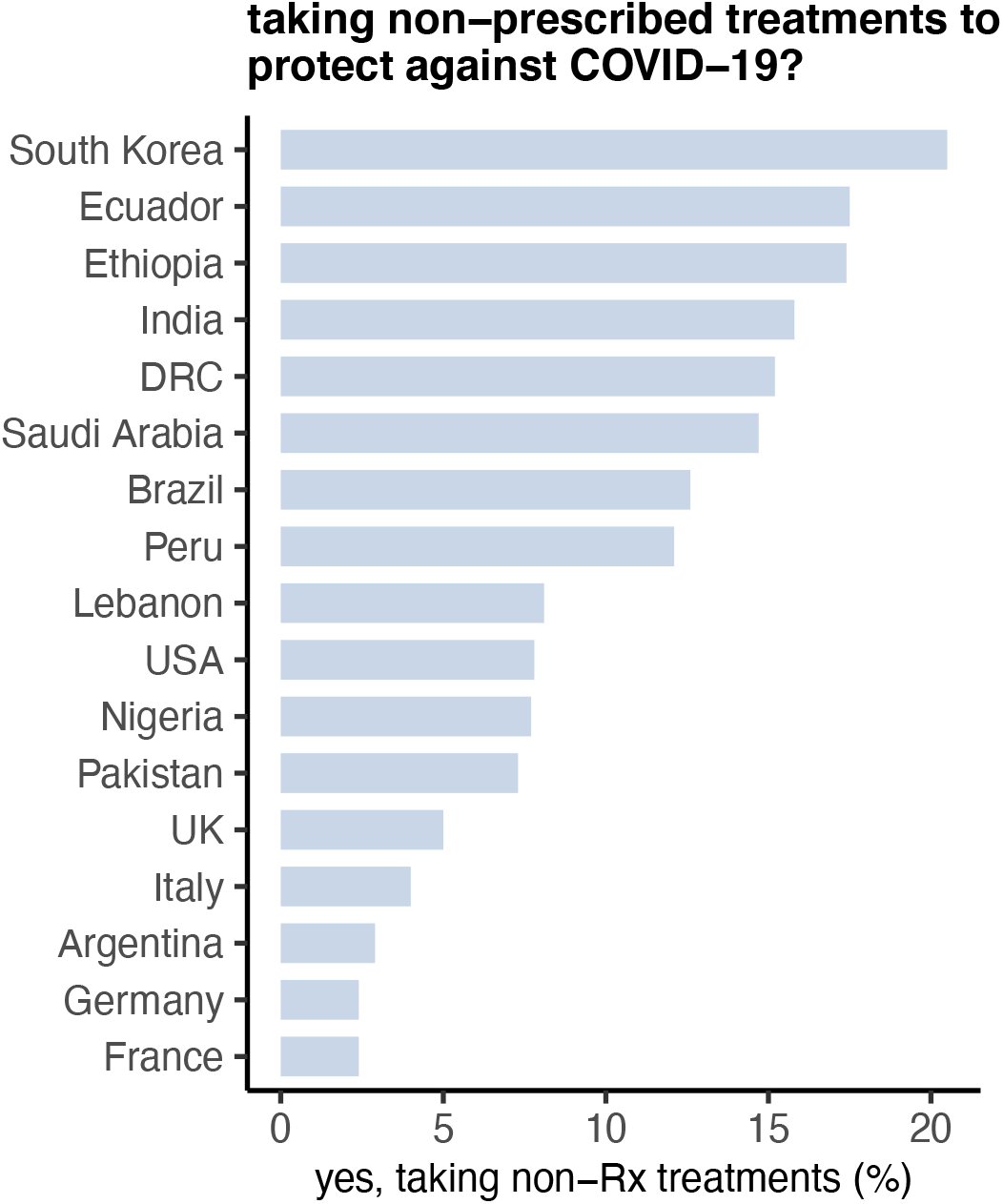
Proportion of respondents in each country reporting that they are taking non-prescribed treatments to protect against COVID-19.

#### Determinants: Emotions

We find strong evidence across multiple countries that individuals who are afraid of catching COVID-19 or who are afraid that someone in their household may catch COVID-19 are far more likely to agree that they would accept a COVID-19 vaccine. Evidence for this effect is found in 12 countries: Argentina (1.85, 1.37 to 2.49), Brazil (1.66, 1.23 to 2.27), Ecuador (1.49, 1.00 to 2.34), France (0.75, 1.31 to 2.33), Germany (1.80, 1.37 to 2.28), Italy (1.40, 1.08 to 1.82), Lebanon (1.11, 0.85 to 1.45), Pakistan (1.64, 1.27 to 2.07), Peru (2.08, 1.40 to 3.04), South Korea (2.02, 1.41 to 2.94), UK (1.84, 1.40 to 2.50), and the USA (1.80, 1.51 to 2.12).

Whether an individual has been feeling fearful in the past few days is also associated with higher uptake intent even after controlling for whether they are afraid that they or someone in their household may catch COVID-19 in Brazil (1.36, 1.01 to 1.82), France (1.85, 1.26 to 2.70), and USA (1.49, 1.49 to 1.86). Feeling fearful in the last few days is also associated with higher uptake intent in Ethiopia (1.45, 1.12 to 1.88) and Nigeria (1.32, 1.03 to 1.67).

Other emotions such as stress, anxiety, and anger appear to be associated with uptake intentions in a small number of countries: stress is associated with increase uptake intent in DRC (1.94, 1.03 to 3.51); increased anger is associated with a decreased uptake intent in Germany (0.60, 0.43 to 0.84); and anxiety is associated with decreased uptake intent in Lebanon (0.73, 0.57 to 0.98), but increased uptake intent in Saudi Arabia (1.26, 1.00 to 1.59).

## Discussion

We conducted a survey of intent to accept a COVID-19 vaccine across 17 countries, as well as potential reasons explaining the variation in acceptance. This study complements three other multi-country studies that have sought to determine barriers to COVID-19 vaccine uptake^6,10,13^.

Objections to vaccination are a global issue, but the level of resistance and the strength of emotion behind them vary considerably^14^. Our results suggest that while socio-demographic factors are associated with COVID-19 vaccine acceptance in a small number of countries (notably, that females were less likely to report intending to accept a COVID-19 vaccine than males in five countries, aligning with recent multi-country evidence^15^), confidence in the safety, effectiveness, and importance of a COVID-19 vaccine and feeling afraid that oneself or a family member may catch SARS-CoV-2 are associated with uptake intent are more consistently associated with vaccine acceptance.

India ranks highest for intention to take a vaccine against COVID-19 and consistently ranks among the most vaccine confident countries globally^16^. By contrast, France, which ranks among the least vaccine confident countriesy^16–18^ has among the lowest willingness to accept a COVID-19 vaccine in this study, alongside the DRC.

The rise of vaccine hesitancy in Europe, particularly France, has worried experts for the last decade^19^. While there were signs of vaccine confidence recovering in across Europe before the pandemic hit ^16^ and immediately following the first reported cases of COVID-19 in February and March 2020^18^, this study shows that there was more hesitancy towards COVID-19 vaccines in many European countries in December 2020, just before the introduction of the first COVID-19 vaccines in Europe.. These confidence trends need to be closely monitored as the vaccines are rolled-out to entire populations^20^, with new virus variants emerging, political disputes over vaccine supplies, and safety concerns around the Oxford-AstraZeneca vaccine leading to temporary suspensions of the vaccine’s roll-out of in multiple European nations. These incidents can further erode confidence and lead to low uptake of a COVID-19 vaccine.

Latin America has one of the highest rates of COVID-19 death in the world^21^. This study identifies demographic groups with lower vaccine confidence and may thus be a focal point for targeted interventions. In Peru, older groups are more likely to state that they would accept a COVID-19 vaccine than younger groups. In order to maximize the effects of herd (community/indirect) immunity, optimal uptake among non-vulnerable groups is also necessary^22^. Religious affiliation is associated with vaccine intent in Argentina, Brazil, and Peru. More specifically, respondents in all three countries who were not part of the dominant Roman Catholic religious group were less likely to report intent to vaccinate. This finding resonates with previous overall vaccine confidence studies^16^ and current concerns of lower COVID-19 vaccine confidence in minority groups^23,24^. In Brazil, where religious intolerance against religions of African roots (i.e. *Umbanda, Candomblé*) is widespread ^25,26^. Our findings highlight the importance of, in South America and elsewhere, tailoring vaccine confidence strategies to minority groups emerging from the COVID-19 pandemic^27^.

Emotional determinants feature strongly in intent to accept a COVID-19 vaccine. While anger was associated with vaccination intent in a small number of countries, in Germany individuals who had recently felt angry were less likely to state intent to accept a COVID-19 vaccine than those who did not report feeling angry recently. Feeling recently anxious is associated with lower uptake intent in Lebanon but increased uptake intent in Saudi Arabia. The same emotions can lead to different outcome. Emotional drivers of COVID-19 acceptance are also context dependent. Emotional determinants of vaccine uptake are situational, and any drivers and outcomes of different emotions need to be considered in perspective.

Misinformation surrounding COVID-19 has provided significant challenges in overcoming false or misleading information^28,29^. A total of 10.1% of respondents surveyed across all countries in this study report self-medicating against COVID-19, taking non-prescribed treatments to protect themselves against COVID-19. While the reason for this could be linked to false or misleading information, emotional factors such as despair^30,31^ could also potentially be driving these outcomes.

The emotional harms of the COVID-19 pandemic as well as their impact on mental health are becoming better understood ^32,33^ and have been discussed elsewhere. Likewise, the role of emotion has also been considered in COVID-19 vaccine communication^34^. Nonetheless, to the best of our knowledge, this is the first attempt to study emotional drivers and determinants of intent to accept a COVID-19 vaccine. We chose to investigate more aversive emotions such as anger, stress, anxiety as these were more likely to show a strong relationship to vaccine acceptance. Future studies should also aim to investigate the role of positive emotions, such as hope^35^, and their impact in vaccine decisions. Additional research could better understand the link between the emotional responses to government pandemic interventions and on actual vaccine uptake, beyond intention. Furthermore, qualitative studies in COVID-19 vaccine confidence are needed local emotional idioms and their relation to vaccine confidence and health outcomes^36^.

There are several study limitations to note. The goal of this study was not to find the most informative set of predictors of uptake, but rather to assess the relative strength of socio-demographic versus emotional determinants and better understand the role of recent emotions – possibly driven by the pandemic or government interventions – and their effect on the willingness to accept a COVID-19 vaccine. There therefore could be a set of questionnaire variables (see appendix) that explain more variance in the outcome than those stated and further research could examine these maximally informative variables. Uptake is also likely going to vary substantially within a country due to local factors and local clustering of demographics^5^, which is outside the scope of this study. Moreover, COVID-19 vaccine acceptance will likely change over time. Sub-national temporal monitoring would be useful to establish local hotspots of non-vaccinators and the demographic and emotional groups who are unlikely to vaccinate Moreover, the robust association found between national level vaccine confidence and intent to accept a COVID-19 vaccine indicates that previous high vaccine confidence could be an indicator of confidence in future COVID vaccines.

#### Evidence before this study

We have previously done three systematic reviews identifying the key determinants of vaccine hesitancy to inform questionnaire design around vaccine confidence. These survey questions have been continually updated by the Vaccine Confidence Project in light of new information around the COVID-19 pandemic. Vaccine refusals have recently contributed to increases in childhood and adult disease outbreaks globally over the past few years, and it is therefore crucial to monitor confidence and vaccine intent of novel COVID-19 vaccines to inform country and cohort-specific intervention strategies to bolster vaccine uptake. We have identified three studies that probe COVID-19 vaccine uptake intent via a nationally representative survey design that interview respondents in more than two countries. (There are, in addition, dozens of country-specific studies that use a variety of surveying techniques). Across the majority of countries investigated to date, males tend to be more likely to state intent to accept a COVID-19 vaccine. Across previous studies, covariates under investigation varied depending on the research question being asked (from misinformation exposure to trust in key sources) complicating large, cross-country comparisons of barriers to uptake.

#### Added value of this study

To the best of our knowledge, this study contains the largest sample size of any multi-country survey to date, with over 19,000 individual responses from 17 countries. To identify key determinants of COVID-19 uptake intent, and to compare these across countries, a standard set of socio-demographic covariates are used: sex, age, highest education level, religious status, and employment status. In addition, the association of recent emotions with vaccine uptake intent and COVID-19 vaccine confidence is considered. These common metrics allow meaningful cross-country comparisons of COVID-19 vaccine sentiment and provide a means to measure future confidence in COVID-19 vaccines and vaccination programmes and to which to assess the success of vaccination policy.

#### Implications of all the available evidence

This study provides novel insights into worldwide variations in COVID-19 vaccine uptake intent and presents the country-dependent factors that may modulate uptake decisions. The study findings are discussed in light of past and ongoing vaccine confidence issues in the 17 countries studied. In light of reported side-effects surrounding some COVID-19 vaccines, a key implication is to highlight the regular monitoring of vaccine confidence levels to identify spatio-temporal trends and changes in sentiment that may suggest the need for policy interventions to sustain or bolster confidence.

## Data Availability

All data used in the study can be provided upon reasonable request to the corresponding author.

## Funding

This project was funded by Janssen Pharmaceutica.

## Role of the funding source

The funders had no role in data collection, questionnaire design, data analysis, data interpretation, or writing of this study. The corresponding author had full access to all the data in the study and had final responsibility for the decision to submit for publication.

## Conflicts of interest

HJL, CS, and AdF are involved in Vaccine Confidence Project collaborative grants with Janssen, GlaxoSmithKline and Merck. HJL has also received other support for participating in Merck meetings and GlaxoSmithKline advisory roundtables. HJL is a member of the Merck Vaccine Confidence Advisory Board.

## Notes

### Funding Statement

This study was funded by Janssen.

### Author Declarations

Ethical approval for this study was granted by the LSHTM Ethics Committee on 15 June 2020 with reference 22130.

