## appendix for "COVID-19 vaccine acceptance and its socio-demographic and emotional determinants: a multi-country cross-sectional study"

### Appendix 1: Additional data

| | | Variable | N | % | Strongly agree | | Agree | | Do not know | | Disagree | | Strongly disagree | |
|---|---|---|---|---|---|---|---|---|---|---|---|---|---|---|
| Socio-econo-demographic factor | sex | female | 9287 | 48.4% | 3539 | 38.1% | 3035 | 32.7% | 1016 | 10.9% | 914 | 9.8% | 784 | 8.4% |
| | | male | 9913 | 51.6% | 4293 | 43.3% | 3406 | 34.4% | 627 | 6.3% | 807 | 8.1% | 780 | 7.9% |
| | age | 18-24 years | 3381 | 17.6% | 1404 | 41.5% | 1167 | 34.5% | 217 | 6.4% | 334 | 9.9% | 259 | 7.7% |
| | | 25-34 years | 4231 | 22.0% | 1776 | 42.0% | 1431 | 33.8% | 250 | 5.9% | 410 | 9.7% | 363 | 8.6% |
| | | 35-44 years | 3526 | 18.4% | 1490 | 42.3% | 1177 | 33.4% | 238 | 6.8% | 317 | 9.0% | 303 | 8.6% |
| | | 45-54 years | 3060 | 15.9% | 1186 | 38.8% | 1014 | 33.1% | 317 | 10.3% | 265 | 8.7% | 278 | 9.1% |
| | | 55-64 years | 2518 | 13.1% | 964 | 38.3% | 845 | 33.6% | 299 | 11.9% | 215 | 8.6% | 194 | 7.7% |
| | | 65+ years | 2485 | 12.9% | 1011 | 40.7% | 806 | 32.4% | 322 | 13.0% | 179 | 7.2% | 167 | 6.7% |
| | education | none/other | 1078 | 5.6% | 451 | 41.8% | 298 | 27.6% | 108 | 10.0% | 104 | 9.6% | 118 | 11.0% |
| | | primary | 2080 | 10.8% | 971 | 46.7% | 631 | 30.3% | 189 | 9.1% | 136 | 6.5% | 154 | 7.4% |
| | | secondary | 7919 | 41.2% | 3121 | 39.4% | 2709 | 34.2% | 744 | 9.4% | 702 | 8.9% | 644 | 8.1% |
| | | undergraduate degree | 6064 | 31.6% | 2344 | 38.6% | 2161 | 35.6% | 485 | 8.0% | 594 | 9.8% | 480 | 7.9% |
| | | postgraduate degree | 2059 | 10.7% | 946 | 45.9% | 643 | 31.2% | 119 | 5.8% | 185 | 9.0% | 167 | 8.1% |
| | work status | full-time | 7687 | 40.0% | 3259 | 42.4% | 2595 | 33.8% | 496 | 6.5% | 662 | 8.6% | 674 | 8.8% |
| | | part-time | 2321 | 12.1% | 898 | 38.7% | 803 | 34.6% | 209 | 9.0% | 221 | 9.5% | 190 | 8.2% |
| | | unemployed | 2420 | 12.6% | 888 | 36.7% | 805 | 33.3% | 227 | 9.4% | 267 | 11.1% | 232 | 9.6% |
| | | retired / disabled | 2493 | 13.0% | 963 | 38.6% | 848 | 34.0% | 346 | 13.9% | 178 | 7.1% | 159 | 6.4% |
| | | student | 1856 | 9.7% | 779 | 42.0% | 678 | 36.5% | 107 | 5.8% | 175 | 9.4% | 117 | 6.3% |
| | | housewife | 2152 | 11.2% | 978 | 45.4% | 635 | 29.5% | 183 | 8.5% | 193 | 8.9% | 163 | 7.6% |
| | | prefer not to say | 272 | 1.4% | 66 | 24.4% | 77 | 28.2% | 76 | 27.9% | 24 | 8.8% | 29 | 10.6% |
| | religious affiliation | Atheist/agnostic | 2158 | 12.1% | 759 | 35.2% | 796 | 36.9% | 230 | 10.7% | 221 | 10.2% | 153 | 7.1% |
| | | Buddhist | 205 | 1.1% | 74 | 36.0% | 89 | 43.4% | 9 | 4.3% | 22 | 10.8% | 11 | 5.5% |
| | | Muslim | 4245 | 23.7% | 1919 | 45.2% | 1478 | 34.8% | 127 | 3.0% | 404 | 9.5% | 316 | 7.4% |
| | | other Christian | 1556 | 8.7% | 474 | 30.4% | 527 | 33.9% | 171 | 11.0% | 184 | 11.8% | 200 | 12.9% |
| | | other religion | 876 | 4.9% | 271 | 30.9% | 282 | 32.2% | 114 | 13.0% | 107 | 12.2% | 102 | 11.7% |
| | | Protestant | 1908 | 10.7% | 681 | 35.7% | 691 | 36.2% | 175 | 9.2% | 176 | 9.2% | 184 | 9.7% |
| | | Roman Catholic | 4468 | 25.0% | 1715 | 38.4% | 1588 | 35.5% | 500 | 11.2% | 344 | 7.7% | 321 | 7.2% |
| | | Russ/Eastern Orthodox | 789 | 4.4% | 409 | 51.8% | 257 | 32.5% | 20 | 2.6% | 63 | 8.0% | 40 | 5.0% |
| | | prefer not to say | 1696 | 9.5% | 516 | 30.4% | 565 | 33.3% | 282 | 16.6% | 167 | 9.8% | 168 | 9.9% |
| | child | No children under 18 | 9993 | 52.0% | 3758 | 37.6% | 3414 | 34.2% | 1075 | 10.8% | 914 | 9.1% | 832 | 8.3% |
| | | Yes, children under 18 | 9207 | 48.0% | 4073 | 44.2% | 3027 | 32.9% | 568 | 6.2% | 807 | 8.8% | 732 | 8.0% |
| vaccine confidence | IMP | agree | 16330 | 85.0% | 7563 | 46.3% | 6045 | 37.0% | 1113 | 6.8% | 1063 | 6.5% | 546 | 3.3% |
| | | disagree | 2871 | 15.0% | 269 | 9.4% | 397 | 13.8% | 531 | 18.5% | 657 | 22.9% | 1018 | 35.5% |
| | SAF | agree | 13241 | 69.0% | 6996 | 52.8% | 5037 | 38.0% | 374 | 2.8% | 559 | 4.2% | 275 | 2.1% |
| | | disagree | 5960 | 31.0% | 835 | 14.0% | 1404 | 23.6% | 1270 | 21.3% | 1161 | 19.5% | 1289 | 21.6% |
| | EFF | agree | 14141 | 73.6% | 7057 | 49.9% | 5368 | 38.0% | 571 | 4.0% | 759 | 5.4% | 386 | 2.7% |
| | | disagree | 5060 | 26.4% | 774 | 15.3% | 1074 | 21.2% | 1072 | 21.2% | 962 | 19.0% | 1178 | 23.3% |
| COVID-19 | at-risk | no | 12840 | 66.9% | 5281 | 41.1% | 4209 | 32.8% | 1004 | 7.8% | 1194 | 9.3% | 1153 | 9.0% |
| | | yes | 5852 | 30.5% | 2434 | 41.6% | 2072 | 35.4% | 516 | 8.8% | 464 | 7.9% | 366 | 6.3% |
| | | prefer not to say | 508 | 2.6% | 117 | 23.0% | 161 | 31.6% | 124 | 24.4% | 62 | 12.2% | 45 | 8.8% |
| | Informed | well | 17378 | 90.5% | 7228 | 41.6% | 5864 | 33.7% | 1426 | 8.2% | 1489 | 8.6% | 1372 | 7.9% |
| | | not well | 1765 | 9.2% | 593 | 33.6% | 562 | 31.9% | 200 | 11.3% | 231 | 13.1% | 179 | 10.1% |
| | | I have never heard of it | 49 | 0.3% | 8 | 16.9% | 10 | 19.4% | 18 | 35.7% | 1 | 2.4% | 13 | 25.6% |
| | | do not know | 8 | 0.0% | 3 | 37.5% | 5 | 62.5% | 0 | 0.0% | 0 | 0.0% | 0 | 0.0% |
| | know some-one | No | 13225 | 68.9% | 5422 | 41.0% | 4371 | 33.0% | 1101 | 8.3% | 1155 | 8.7% | 1176 | 8.9% |
| | | Yes | 5976 | 31.1% | 2409 | 40.3% | 2071 | 34.6% | 542 | 9.1% | 565 | 9.5% | 388 | 6.5% |
| | | do not know / refused | 5 | 0.0% | 3 | 58.0% | 1 | 19.2% | 0 | 0.0% | 1 | 14.7% | 0 | 8.1% |
| | non-Rx | No | 17259 | 89.9% | 6917 | 40.1% | 5842 | 33.9% | 1546 | 9.0% | 1550 | 9.0% | 1404 | 8.1% |
| | | Yes. What? | 1937 | 10.1% | 911 | 47.1% | 598 | 30.9% | 98 | 5.1% | 169 | 8.8% | 159 | 8.2% |
| Emotions | afraid | Agree | 13673 | 71.2% | 5898 | 43.1% | 4758 | 34.8% | 1193 | 8.7% | 1057 | 7.7% | 767 | 5.6% |
| | | Disagree | 5527 | 28.8% | 1934 | 35.0% | 1683 | 30.4% | 450 | 8.1% | 664 | 12.0% | 797 | 14.4% |
| | stress | No | 12291 | 64.0% | 4974 | 40.5% | 4097 | 33.3% | 1080 | 8.8% | 1091 | 8.9% | 1049 | 8.5% |
| | | Yes | 6910 | 36.0% | 2858 | 41.4% | 2344 | 33.9% | 564 | 8.2% | 630 | 9.1% | 514 | 7.4% |
| | anxiety | No | 13047 | 68.0% | 5237 | 40.1% | 4296 | 32.9% | 1122 | 8.6% | 1211 | 9.3% | 1181 | 9.1% |
| | | Yes | 6153 | 32.0% | 2595 | 42.2% | 2145 | 34.9% | 521 | 8.5% | 510 | 8.3% | 383 | 6.2% |
| | anger | No | 14350 | 74.7% | 5859 | 40.8% | 4863 | 33.9% | 1276 | 8.9% | 1256 | 8.8% | 1097 | 7.6% |
| | | Yes | 4850 | 25.3% | 1973 | 40.7% | 1578 | 32.5% | 368 | 7.6% | 464 | 9.6% | 467 | 9.6% |
| | fear | No | 14396 | 75.0% | 5585 | 38.8% | 4863 | 33.8% | 1334 | 9.3% | 1357 | 9.4% | 1257 | 8.7% |
| | | Yes | 4804 | 25.0% | 2246 | 46.8% | 1578 | 32.8% | 310 | 6.4% | 363 | 7.6% | 307 | 6.4% |

*Table A1. Cross-table of the response variable (vaccination intent) by each level of each explanatory variable*

| *Country* | Methodology | Fieldwork dates |
|---|---|---|
| Argentina | CATI | 16-28 June |
| Brazil | CATI | 16 June - 23 July |
| Democratic Republic of Congo | CATI | 19-28 June |
| Ecuador | CATI | 16 June - 22 July |
| Ethiopia | CATI | 23 June -4 July |
| France | Online | 10-16 June 2020 |
| Germany | Online | 10-16 June 2020 |
| India | CATI | 23 June - 2 July |
| Italy | Online | 10-16 June 2020 |
| Lebanon | CATI | 24 June - 16 July |
| Nigeria | F2F | 15 - 28 June |
| Pakistan | CATI | 12 June - 6 July |
| Peru | CATI | 16 June - 22 July |
| Saudi Arabia | CATI | 25 June - 8 July |
| South Korea | Online | 15-18 June 2020 |
| UK | Online | 5-13 June 2020 |
| USA | Online | 5-15 June 2020 |

***Table A2.*** **Survey fieldwork dates and survey methodology**

#### Appendix 2: Questionnaire

**COVID 19 PERCEPTIONS**
**Spring 2020**
**Master Questionnaire**

**EACH SURVEY WILL BE CARRIED OUT AMONG A NATIONALLY REPRESENTATIVE SAMPLE OF N=1000 ADULTS PER COUNTRY. THE METHODOLOGY (E.G. ONLINE v TELEPHONE v FACE-TO-FACE) WILL BE DETERMINED BY THE COUNTRY**

*SECTION 1 : AWARENESS OF COVID*

Q1 Firstly I would like to ask you how well informed you feel you are about Coronavirus (COVID19)?

1 Very well informed
2 Fairly well informed
3 Not very well informed
4 Not informed at all
5 I have never heard of it (do not read out)

Q2 Do you personally know anyone who has tested positive for COVID-19? If yes, was that a family member, a work colleague, a friend or someone else? (Code all mentions)

1 No
2 Yes – myself
3 Yes – family member in my household
4 Yes – family member outside my household
5 Yes – a work colleague
6 Yes – a friend
7 Yes – Someone else.

Q3 What are the three sources you trust the most for information about Coronavirus (COVID-19)? (Script to ensure we have $1^{st}$,$2^{nd}$ and $3^{rd}$).

1 National television
2 Satellite/international television channels
3 Radio
4 Newspapers
5 Social media (such as Facebook, Twitter etc)
6 WhatsApp
7 National public health authorities (e.g. medical authorities/doctors)
8 International health authorities (e.g. WHO)
9 Government websites specifically
10 Other government communications online (e.g. social media)
11 The internet or search engines (e.g. Google)
12 Family and friends
13 Work/school/college
14 Other (specify)____________
15 I don't trust any sources (Do not read out)

*SECTION 2 : FEELINGS, BEHAVIOUR AND OPINION*

Q4 Which of the following feelings and emotions have you experienced in the last few days? Please indicate all those that you have experienced.

ROTATE ORDER

| | | | |
|---|---|---|---|
| Happiness | Trust | Stress | Hope |
| Anxiety | Positivity | Dynamism | Depression |
| Boredom | Anger | Nervousness | Concern |
| Fear | Sadness | Fatigue | Uncertainty |
| Fun | | | |

98 None of the above

Q5 And more generally, on a scale of 0 to 10 how would you describe your morale over the last 7 days? Where 0 is extremely low morale and 10 is extremely high

0 1 2 3 4 5 6 7 8 9 10

Q6 How regularly are you doing each of the following compared with three months ago?

| ROTATE ORDER | A lot more regularly | A little more regularly | About the same | Less regularly | Do not kno[illegible] |
|---|---|---|---|---|---|
| Wearing a face mask (or appropriate substitute for example a scarf) | 1 | 2 | 3 | 4 | 9 |
| Washing my hands | 1 | 2 | 3 | 4 | 9 |
| Covering my nose or mouth when sneezing/coughing | 1 | 2 | 3 | 4 | 9 |
| Going food shopping | 1 | 2 | 3 | 4 | 9 |
| Going out in general | 1 | 2 | 3 | 4 | 9 |
| Using public transport | 1 | 2 | 3 | 4 | 9 |
| Having guests in your house | 1 | 2 | 3 | 4 | 9 |
| Gathering socially with small groups of friends | 1 | 2 | 3 | 4 | 9 |

Q7a Thinking about yesterday, about how many times would you say you washed your hands with soap and water?

/___/ Number of times [SCRIPTER: MAXIMUM 100]

Q7b And again, thinking about yesterday, about how many times would you say you used hand sanitizer?

/___/ Number of times [SCRIPTER: MAXIMUM 100]

Q8 Are you taking any non prescribed medicines or treatments that you have read/heard about that are said to help protect yourself specifically against Coronavirus (COVID-19)? By non-prescribed, I mean over the counter medicine, herbal medicine, alternative treatments or supplements **purchased specifically to protect you from catching Coronavirus (COVID 19)**

1 Yes. What?________________________
2 No

Q9 How strongly do you agree or disagree with the following statements

| ROTATE ORDER | Strongly agree | Agree | Disagree | Strongly disagree | Do not kno |
|---|---|---|---|---|---|
| I am afraid that either myself or someone in my household may catch Coronavirus (COVID-19) | 1 | 2 | 3 | 4 | 9 |
| I think the (INSERT COUNTRY) Government is handling the Coronavirus pandemic well | 1 | 2 | 3 | 4 | 9 |
| Doctors and hospitals are doing the best they can to manage cases and save lives | 1 | 2 | 3 | 4 | 9 |
| The police have intervened too much in public life in regards to the restrictions put in place relating to COVID-19 | 1 | 2 | 3 | 4 | 9 |

Q10a On a scale of -10 to +10, where -10 means you think public health should be totally prioritised right now and +10 means you think the economy should be totally prioritised, where do you believe the Government's immediate focus should be?

Q10b And where do you think the Government is on this same scale right now?

Public health Economy

| -10 | -9 | -8 | -7 | -6 | -5 | -4- | -3 | -2 | -1 | 0 | 1 | 2 | 3 | 4 | 5 | 6 | 7 | 8 | 9 | 10 |
|---|---|---|---|---|---|---|---|---|---|---|---|---|---|---|---|---|---|---|---|---|

*SECTION 3: VACCINES*

Q11 Imagine a new vaccine for Coronavirus (COVID-19) was fast tracked and approved by medical professionals and regulators. How strongly do you agree or disagree with the following statements?

| ROTATE ORDER | Strongly agree | Agree | Disagree | Strongly disagree | Do n know |
|---|---|---|---|---|---|

| | | | | | |
|---|---|---|---|---|---|
| If a new Coronavirus (COVID-19) vaccine became publicly available I would take it. | 1 | 2 | 3 | 4 | 9 |
| I think a new Coronavirus (COVID 19) vaccine would be safe | 1 | 2 | 3 | 4 | 9 |
| I think a new Coronavirus (COVID 19) vaccine would be important | 1 | 2 | 3 | 4 | 9 |
| I think a new Coronavirus (COVID 19) vaccine would be effective | 1 | 2 | 3 | 4 | 9 |
| The country in which the vaccine was manufactured would have no impact on my uptake | 1 | 2 | 3 | 4 | 9 |

FOR THOSE WHO DISAGREE TO TAKE THE VACCINE [IF Coded 3 or 4 at Q11 statement 1]

Q11b Why would you not be willing to take a vaccine against Coronavirus (COVID-19). PROBE. And why else?

PLEASE WRITE IN ________________________________________

(CODE FRAME FOR ANALYSIS. CAWI DO NOT SHOW)

1 I do not feel I am at risk of catching the virus
2 I am confident there will be other effective treatments soon
3 I do not yet know enough about the vaccine to make a decision
4 Approval/Development for the vaccine may be rushed and it may not be thoroughly tested
5 I believe vaccines can give you the disease they are designed to protect you against
6 OTHER CODES
7 I Do not know

Q12 Assuming a COVID-19 vaccination was approved and manufactured, which of the following groups, if any, do you think should be prioritised for the vaccine? PLEASE TICK UP TO THREE RESPONSES

1 Healthcare workers (eg hospital staff, care workers etc)
2 Other key workers (eg teachers, shopkeepers, delivery people etc)
3 The elderly in general
4 Those living in care homes
5 Vulnerable populations (eg those with underlying health conditions such as lung conditions, heart disease, a weakened immune system etc.)
6 Those unable to follow social distancing rules due to living conditions (e.g. living in a crowded setting, need to go out to make a living)
7 Children
8 All adults
9 Other (specify)__________
10 NO priority – it should be open to all.
11 Don't know

Q13 How strongly do you agree or disagree with each of the following statements about <u>vaccines in general</u>?

| ***Rotate Statements*** | Strongly agree | Tend to agree | Tend to disagree | Strongly disagree | Do not know/ NR**(Online: Show)** |
|---|---|---|---|---|---|
| a) Vaccines are important for people of all ages to have | **1** | **2** | **3** | **4** | **5** |
| b) Vaccines are important for children to have | **1** | **2** | **3** | **4** | **5** |
| c) Overall I think vaccines are safe | **1** | **2** | **3** | **4** | **5** |

| | | | | | |
|---|---|---|---|---|---|
| d) Overall I think vaccines are effective | **1** | **2** | **3** | **4** | **5** |
| e) Vaccines are compatible with my religious beliefs | **1** | **2** | **3** | **4** | **5** |

**Q14** How strongly do you agree or disagree with each of the following statements about the <u>MMR and seasonal influenza vaccine</u>?

| | | | | | |
|---|---|---|---|---|---|
| a) Overall I think the MMR vaccine is safe | **1** | **2** | **3** | **4** | **5** |
| b) Overall I think the seasonal influenza vaccine is safe | **1** | **2** | **3** | **4** | **5** |
| c) I think the MMR vaccine is important for children to have | **1** | **2** | **3** | **4** | **5** |
| d) I think the seasonal influenza vaccine is important | **1** | **2** | **3** | **4** | **5** |
| e) Overall I think the MMR vaccine is effective | **1** | **2** | **3** | **4** | **5** |
| f) Overall I think the seasonal influenza vaccine is effective | **1** | **2** | **3** | **4** | **5** |

ROTATE ORDER OF STATEMENTS A and B

Q15a When considering your willingness to vaccinate yourself in general, would you say the global Coronavirus (COVID19) pandemic has made you now much less likely, somewhat less likely, somewhat more likely or a lot more likely to vaccinate compared with one year ago – or perhaps there has been no change in your views to vaccines?

Q15b According to scientific evidence, on average a person with coronavirus will infect three individuals. On average a person with measles will infect 14 others. Does this make you much less likely, somewhat less likely, somewhat more likely or a lot more likely to support measles-containing vaccinations – or perhaps this has no change in your views to vaccines?

| ***Rotate Statements*** | A lot more likely | Somewhat more likely | Somewhat less likely | A lot less likely | There has been no change |
|---|---|---|---|---|---|
| Statement 15a | **1** | **2** | **3** | **4** | **5** |
| Statement 15b | **1** | **2** | **3** | **4** | **5** |

FOR COUNTRIES WITH EXPERIENCE OF PREVIOUS PANDEMICS ONLY, I.E. South Korea (SARS), Saudi Arabia (MERS) and DRC (Ebola).

Q16. Thinking about the previous pandemic in your country, i.e. [INSERT COUNTRY-SPECIFIC PANDEMIC], how strongly do you agree or disagree with the following statements? As a result of my experience with (SARS/MERS/EBOLA)…

| ***Rotate Statements*** | Strongl y agree | Tend to agree | Tend to disagre e | Strongl y disagre e | Do not know/ NR**(Onl** |
|---|---|---|---|---|---|

| | | | | | ine:<br>Show) |
|---|---|---|---|---|---|
| a) I know what precautions I need to take as an individual to protect myself and others against coronavirus. | **1** | **2** | **3** | **4** | **5** |
| b) I have **more** confidence in our government's ability to deal with COVID-19 effectively. | **1** | **2** | **3** | **4** | **5** |
| c) our country is **better** prepared to deal with COVID-19. | **1** | **2** | **3** | **4** | **5** |
| d) our country will be **better** prepared with the economic and social consequences of the outbreak in the long-term | **1** | **2** | **3** | **4** | **5** |

**DEMOGRAPHICS**

**Now just a few more questions about yourself to ensure that we have a representative sample**

**D1** Gender

Male ............................................................................................................ 1
Female ......................................................................................................... 2

**D2** What is your age?

________ years

**D2_CODE** Age Recode

18-24 years ................................................................................................... 1
25-34 years ................................................................................................... 2
35-44 years ................................................................................................... 3
45-54 years ................................................................................................... 4
55-64 years ................................................................................................... 5
65+ years .......................................................................................................

**D3** What is the highest level of education you completed?

None/no formal education .............................................................................. 1
Primary ........................................................................................................ 2
Secondary .................................................................................................... 3
University ..................................................................................................... 4
Master/PhD ................................................................................................... 5
Other education level (specify)______________................................................... 6
*[categories to be customised to country.]*

**D4** Do you have any children under 18 years of age living in your household, and if yes how old are they?

None............................................................................................................ 1
Children up to the age of 2 years .................................................................... 2

Children of 3 to 6 years ........................................ 3
Children of 7 to 12 years ........................................ 4
Children of 13 to 17 years ........................................ 5

**D5** Do you or does anyone in your household have underlying health conditions which may increase their risk from coronavirus? (Conditions such as lung conditions, pregnancy, heart disease, a weakened immune system etc.)

Yes ........................................ 1
No ........................................ 2
Prefer not to say ........................................ 98

**D6.** Which of the following best describes your working status?

Working full-time (include self-employed) ........................................ 1
Working Part-time ........................................ 2
Unemployed ........................................ 3
Student ........................................ 4
Housewife ........................................ 5
Retired/Disabled ........................................ 6
Refused/DNK/DNA ........................................ 9

**D7.** Which sort of town or village do you live in?

Capital ........................................ 1
Big city ........................................ 2
Small city ........................................ 3
Village ........................................ 4
Refused/ DNA ........................................ 9

**D8** Do you consider yourself:

Roman Catholic ........................................ 01
Russian or Eastern Orthodox ........................................ 02
Protestant ........................................ 03
Other Christian ........................................ 04
Hindu ........................................ 05
Muslim ........................................ 06
Jewish ........................................ 07
Buddhist ........................................ 08
Other ........................................ 09
Atheist/agnostic ........................................ 10
Refused/DNK/DN
